# Viral loads and profile of the patients infected with SARS-CoV-2 Delta, Alpha or R.1 variants in Tokyo

**DOI:** 10.1101/2021.09.21.21263879

**Authors:** Chihiro Tani-Sassa, Yumi Iwasaki, Naoya Ichimura, Katsutoshi Nagano, Yuna Takatsuki, Sonoka Yuasa, Yuta Takahashi, Jun Nakajima, Kazunari Sonobe, Yoko Nukui, Hiroaki Takeuchi, Kousuke Tanimoto, Yukie Tanaka, Akinori Kimura, Shuji Tohda

**Affiliations:** Clinical Laboratory, Tokyo Medical and Dental University (TMDU) Hospital, Tokyo, Japan; Infection Control and Prevention, Tokyo Medical and Dental University (TMDU) Hospital; Department of Molecular Virology, Tokyo Medical and Dental University (TMDU); Genome Laboratory, Medical Research Institute, Tokyo Medical and Dental University (TMDU); Research Core, Institute of Research, Tokyo Medical and Dental University (TMDU); Institute of Research, Tokyo Medical and Dental University (TMDU)

**Keywords:** SARS-CoV-2, COVID-19, Delta variant, viral load, PCR

## Abstract

The rapid spread of the Delta variant of SARS-CoV-2 became a serious concern worldwide in summer 2021. We examined the copy number and variant types of all SARS-CoV-2-positive patients who visited our hospital from February to August 2021 using PCR tests. Whole genome sequencing was performed for some samples. The R.1 variant (B.1.1.316) was responsible for most infections in March, replacing the previous variant (B.1.1.214); the Alpha (B.1.1.7) variant caused most infections in April and May; and the Delta variant (B.1.617.2) was the most prevalent in July and August. There was no significant difference in copy numbers among the previous variant cases (n=29, median 3.0×10^4^ copies/μL), R.1 variant cases (n=28, 2.1×10^5^ copies/μL), Alpha variant cases (n=125, 4.1×10^5^ copies/μL), and Delta variant cases (n=106, 2.4×10^5^ copies/μL). Patients with Delta variant infection were significantly younger than those infected with R.1 and the previous variants, possibly because many elderly individuals in Tokyo were vaccinated between May and August. There was no significant difference in mortality among the four groups. Our results suggest that the increased infectivity of Delta variant may be caused by factors other than the higher viral loads. Clarifying these factors is important to control the spread of Delta variant infection.

## INTRODUCTION

The rapid spread of the Delta variant (B.1.617.2) of SARS-CoV-2 became a serious problem worldwide in summer 2021 (1). A precise understanding of the features of the Delta variant is crucial for controlling its spread. Tokyo Medical and Dental University Hospital is located in the center of Tokyo and mainly treats patients with severe coronavirus disease 2019 (COVID-19) from all over Tokyo. We examined the copy number and variant types of the samples from all COVID-19 patients who visited our hospital.

We previously reported that the R.1 variant (the sublineage of B.1.1.316) rapidly replaced the previously existing strain, B.1.1.214 (originating from the European lineage), in Tokyo in March 2021 (2). After that, the Alpha variant (B.1.1.7) and the Delta variant prevailed.

Information on SARS-CoV-2 variants and patient numbers is provided by the Tokyo Metropolitan Institute of Public Health (3), but is based on a sampling survey and does not include patient data such as viral loads. In this paper, we show the precise viral loads and patient profiles based on our clinical data.

## MATERIALS AND METHODS

### Patients and samples

Data was collected from COVID-19 patients whose diagnoses were confirmed by PCR tests performed from February 1 to August 31, 2021. Nasopharyngeal swab samples were obtained at the time of outpatient visit or admission. The samples were immersed in test tubes containing 1 mL of phosphate-buffered saline containing 1% dithiothreitol prior to analysis.

This study was approved by the Medical Research Ethics Committee of Tokyo Medical and Dental University (approval number: M2020-004) and was conducted in accordance with the ethical standards of the 1964 Helsinki Declaration.

### Quantitative PCR test

To detect and quantify SARS-CoV-2 in the samples, one-step reverse transcription-quantitative PCR was performed without viral RNA purification using the 2019 Novel Coronavirus Detection Kit (Shimadzu Corp., Kyoto, Japan) and the QuantStudio 5 Dx Real-Time PCR System (Thermo Fisher Scientific, Inc., Waltham, MA, USA). The copy numbers were estimated from the calibration lines of the concentration-known standard samples.

### Variant screening PCR test

To determine the SARS-CoV-2 variant for each patient, we used melting curve analysis of PCR products. Viral RNA was purified from the swab samples that were positive for SARS-CoV-2, using the EZ1 Virus Mini Kit v2.0 and EZ1 advanced XL (QIAGEN; Venlo, Netherlands). RT-PCR was performed using the LightCycler Multiplex RNA Virus Master (Roche Molecular Systems, Inc.; Basel, Switzerland) and primers and probes provided in the VirSNiP SARS-CoV-2 kits for Spike N501Y, E484K, and L452R (TIB Molbiol; Berlin, Germany). The melting curve analysis of the PCR products was performed according to the manufacturer’s instructions. The 501N, 501Y, 484E, 484K, 484Q, 452R, and 452L types were determined by their melting temperature profiles.

Whole genome sequencing (WGS) was performed to identify the lineages of representative samples using a next-generation sequencer. Libraries were prepared using the QIAseq SARS-CoV-2 Primer Panel Kit (QIAGEN) and sequenced using the MiSeq (Illumina, Inc.; San Diego, USA). Alignment and variant calling were performed using CLC Genomics Workbench software (QIAGEN).

### Statistical analysis

The difference in copy numbers and ages among the four types of variant cases was evaluated using the Steel-Dwass test. The difference in the rate of ICU admission and mortality was evaluated using the chi-square test.

## RESULTS

Based on the results of variant screening PCR, we divided the samples into four groups, as shown in Table 1. WGS was conducted on approximately one-fifth of samples for each variant group, these results consistently agreed with our grouping, confirming that our process was appropriate. PANGO lineages (4) were determined using the WGS data from the representative samples. The strains identified as 501Y and 484K (Beta or Gamma variant) or 484Q and 452L (Kappa variant) were not found in our study sample.

**Table 1.** Outline of each variant type

| Mutation | 501 | 484 | 452 | PANGO lineage |
| --- | --- | --- | --- | --- |
| Previous strain | N | E | L | B.1.1.214 |
| R.1 | N | K | L | B.1.1.316 |
| Alpha | Y | E | L | B.1.1.7 |
| Delta | N | E | R | B.1.617.2 |

The PANGO lineage was determined by whole genome sequencing of representative samples from each group.

The sequential transitions of the variants are shown in Fig. 1. The R.1 variant replaced the previous variants in March, as previously reported (2). The Alpha variant represented the majority of infections in April and May. The first Delta variant case in our hospital was identified in mid-May and the Delta variant was predominant in July and August.

**Fig. 1.**
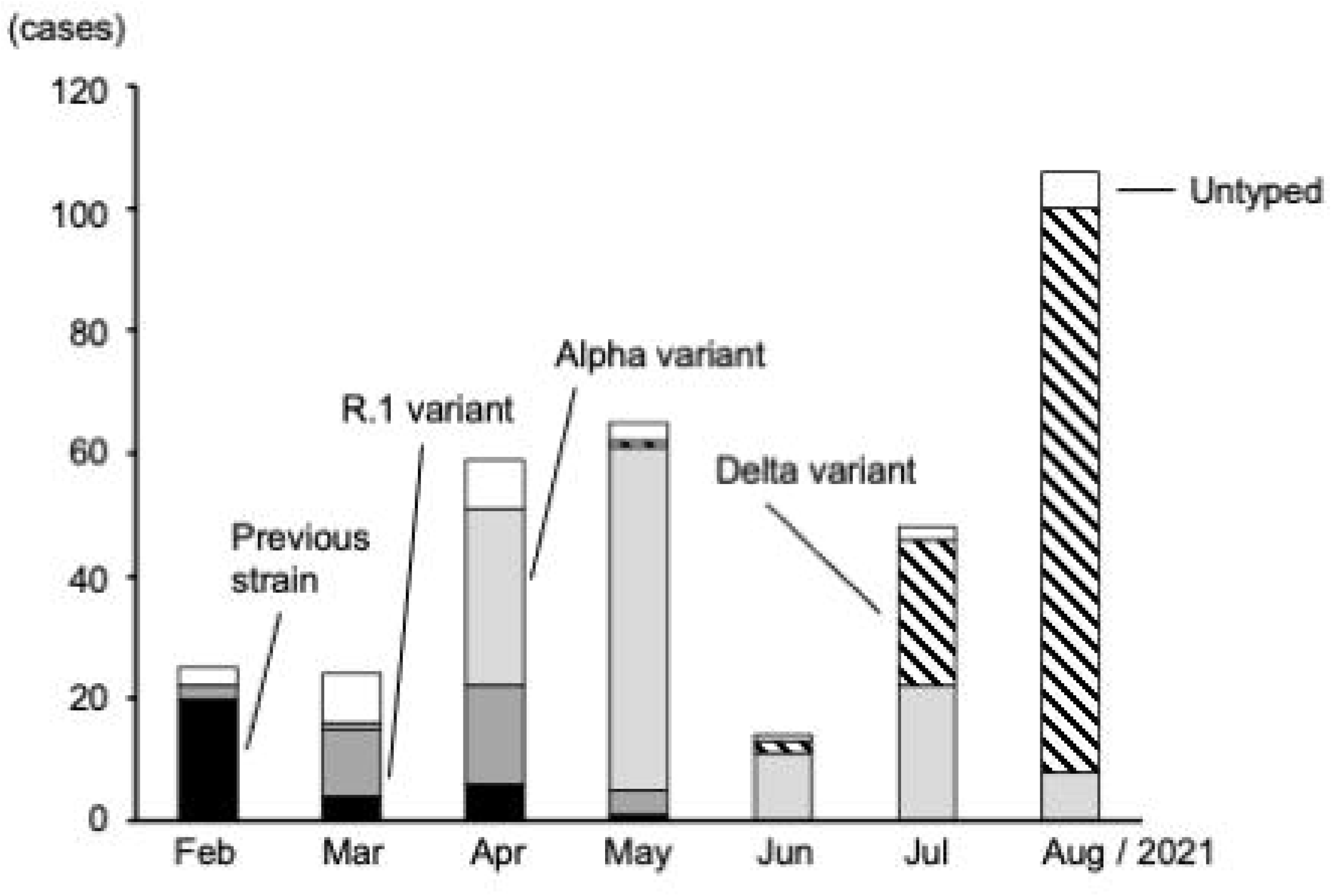
Sequential transition of four types of variant cases determined by PCR-based melting curve analysis. Untyped bar represents the samples of which variant type was not determined because of that the PCR products were not generated probably due to small copy number.

The variant types from 20 samples could not be determined as variant screening PCR did not produce any PCR products; this is probably because the samples had a very small copy number (data not shown). Untyped samples (shown as Untyped bar in Fig. 1) were excluded from the following analysis.

The clinical profiles of the 288 patients enrolled are shown in Table 2. Comparing the age of each variant group, there was no significant difference between Delta and Alpha cases. However, the Delta variant cases were significantly younger than the R.1 variant cases (p= 5 × 10^−5^) and the previous strain cases (p= 6 × 10^−7^).

**Table 2.** Clinical profiles of patients with four different variants of SARS-CoV-19

| **Variant type** | **Previous** | **R.1** | **Alpha** | **Delta** |
| --- | --- | --- | --- | --- |
|  | (n=29) | (n=28) | (n=125) | (n=106) |
| Age (years) mean ± S.D. | 68.5 ± 13.8 | 66.9±19.7 | 51.5±15.8 | 47.6±17.6 |
| Male ratio (%) | 70.0 | 57.1 | 72.0 | 64.2 |
| Mortality (number, %) | n=4, 13.8% | n=4, 4.3% | n=4, 3.2% | n=3, 2.8% |
| ICU admission (%) | 44.8 | 25.0 | 22.4 | 17.0 |
| Viral loads (copies/μL)<br>median | $3.0 \times 10^4$ | $2.1 \times 10^5$ | $4.1 \times 10^5$ | $2.4 \times 10^5$ |

As an indicator of increased severity, the rates of admission to the intensive care unit (ICU) were compared among the four groups. The rate of ICU admission for previous strain cases was significantly higher than that of Delta variant cases (p=0.022). There were no significant differences among the other variant pairs. With regard to mortality, no significant difference was found between any pair of groups. The distribution of viral loads as copy numbers in the swab-soaked samples is shown in Fig. 2. There was no significant difference in copy number among the four variant groups. We could not find a clear relationship between copy number and severity (Fig. 2 – upper panel) and between copy number and generation (Fig. 2 – lower panel). The copy numbers of patients who died in the Delta variant group seemed high compared to the numbers of those who did not die, however, this finding could not be confirmed as only three patients died in this group.

**Fig. 2.**
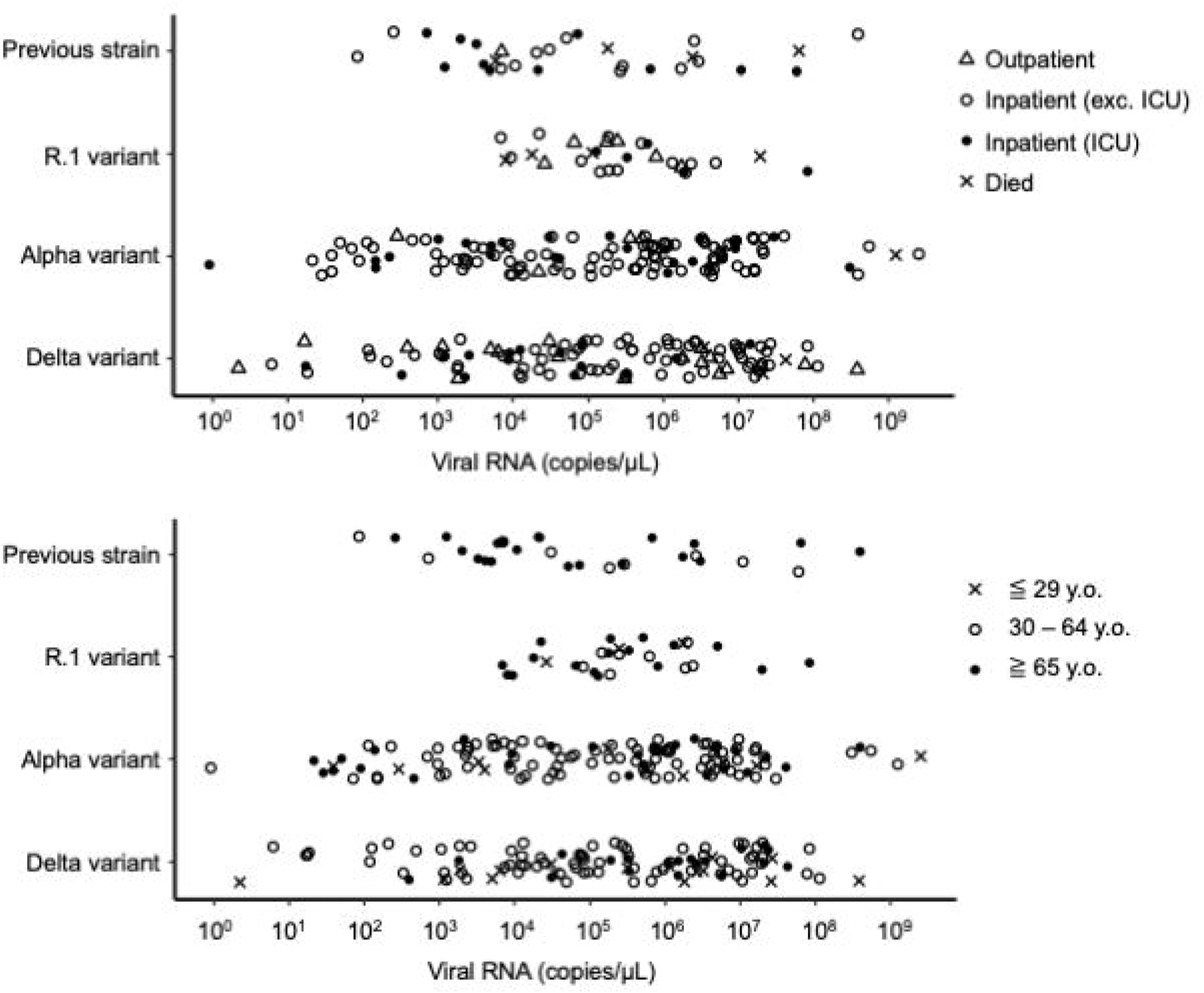
Copy numbers of viral RNA in swab-soaked samples for four types of SARS-CoV-2 variant cases, estimated by reverse transcription-quantitative PCR. Cases are marked according to the severity (upper panel) and the generation (lower panel).

## DISCUSSION

The Delta variant has attracted attention for its increased transmissibility (1). Viral load is thought to be one of the factors that causes increased transmissibility. Li et al. reported that the viral load of Delta variants was more than a thousand times higher than that of the Wuhan strain in the initial wave of 2020 in China (5). Ong et al. also reported that the Delta variant was associated with lower PCR cycle threshold values compared to the wild-type in Singapore (6). Because we had no patients with the Wuhan strains in early 2020, we could not compare the viral load between the delta variant cases and the Wuhan strain cases.

Contrary to these reports, our data showed that the viral loads of the Delta variant cases were no higher than those of the other variant cases. It should be noted that our data may be biased because our hospital specializes in patients with severe disease and therefore may not reflect the distribution of all cases in Tokyo. However, as this policy did not change over the study period, it is unlikely that it affected the distribution of our data.

The Delta variant was reported to be more frequently detected in the younger generation than other variants (7), and our data also showed the same tendency. However, it is speculated that this was not due to the nature of Delta variants, but rather to the fact that many elderly individuals (over the age of 65) were vaccinated from May to August in Tokyo, reducing the infection rates of the elderly.

As an indicator of severity, we focused on the rates of ICU admission and mortality. We showed that the Delta variant did not lead to increased severity compared to the other variants. However, these data might be affected by the selection bias of the patients mentioned above. Our data included small number of outpatients who did not need hospitalization due to mild symptoms, and some patients were transferred to our hospital due to deterioration from other hospitals after several days of hospitalization. This process meant that more than one week may have passed since the onset of illness, which may have affected viral loads in the samples taken at the time of admission to our hospital. In addition, the relatively low mortality of the Delta variant group could be an effect of vaccination in the elderly.

Our results suggest that the increased infectivity of the Delta variant may not be caused by higher viral loads, but by other factors, such as a higher affinity of the mutated spike proteins to the cells (8). Clarifying the mechanism is a future task to control the spread of the Delta variant.

## Data Availability

The data and protocol of RT‐PCR are available from the corresponding author upon request.

## ACKNOWLEDGMENTS

WGS was supported by grant JPMJCR20H2 from JST-CREST and grant 20nk0101612h0901 from the Japan Agency for Medical Research and Development. We would like to thank all the staff involved in COVID-19 treatment.

